# AD plasma biomarkers are stable for an extended period at –20°C: implications for resource-constrained environments

**DOI:** 10.1101/2024.07.17.24310504

**Authors:** Biniyam A. Ayele, Patrice L. Whitehead, Julianna Pascual, Tianjie Gu, Jamie Arvizu, Charles G. Golightly, Larry D. Adams, Margaret A. Pericak-Vance, Jeffery M. Vance, Anthony J. Griswold

## Abstract

Standard procedures for measuring Alzheimer’s disease (AD) plasma biomarkers include storage at -80°C. This is challenging in countries lacking research infrastructure, such -80°C freezer. To investigate stability of AD biomarkers from plasma stored at -20°C, we compared aliquots stored at -80°C and others at -20°C for two, four, six, fifteen, and thirty-five weeks. pTau181, Aβ42, Aβ40, NfL, and GFAP were measured for each timepoint. pTau181 and Aβ42/Aβ40 ratios showed minimal variation for up to 15 weeks. NfL and GFAP had higher variability. This finding of 15-week stability at -20°C enables greater participation in AD biomarker studies in resource constrained environments.

## INTRODUCTION

Globally, Alzheimer’s disease (AD) is the most common cause of dementia accounting for 60%-80% of all cases [1–4]. Over 60% of AD patients reside in low- and middle-income countries (LMIC) with exponential increase in prevalence expected by mid-century [1–4]. AD is characterized by the extracellular deposition of beta amyloid plaques and intracellular deposition of neurofibrillary tau tangles, which gradually result in damage and loss of neuronal cells [5,6]. Current gold standard diagnostic methods to confirm the presence of amyloid beta and tau deposition in the brain are advanced imaging techniques such as amyloid-PET and tau-PET scans or measurement of cerebrospinal fluid (CSF) based biomarkers such as amyloid-β or phosphorylated Tau [7–10]. However, given the invasive and expensive nature of these tests, their use in resource constrained settings is prohibitive. However, blood-based biomarkers (BBM) show potential for aiding in the early and accurate detection of AD neuropathology and identifying presymptomatic or mild cognitive impairment (MCI) stages of AD cases [11–15]. Thus, plasma biomarkers could offer cost-effective, easily accessible, and scalable AD diagnostic options for widespread use in resource constrained settings [7,16].

While plasma isolation is minimally invasive and low cost, appropriate preanalytical handling is critical to limit variability in measurements. As recommended by the Standardization of Alzheimer’s Blood Biomarkers (SABB) working group of the Alzheimer’s Association, plasma should be stored immediately at -20°C for a maximum of two weeks with longer term storage at -80°C [17]. Such a requirement is challenging in low-resource settings where -80°C storage is not readily available nor is shipping of frozen aliquots practical or cost effective. An example of this challenge presents itself in the DAWN study, a multi-center project funded by the National Institute on Aging (NIA). DAWN aims to explore AD in diverse ancestral groups including nine countries from sub-Saharan Africa (SSA) through the Africa Dementia Consortium (AfDC) [18]. This involves collection of detailed clinical and neuropsychological data and blood samples from 5000 indigenous Africans for genomic and biomarkers investigation. Critically, of the nine countries participating in the DAWN study from Africa, only three have local access to -80°C freezers, though all have access to -20°C storage.

While previous studies have indicated that intermittent storage of plasma samples at - 20°C for 2 weeks does not change biomarker levels [17], no study has shown stability of BBM beyond two weeks. Herein we investigate the stability of biomarker measures of phosphorylated threonine-181 of Tau (pTau181), amyloid-β 40 & 42 (Aβ40 and Aβ42), neurofilament light chain (NfL), and glial fibrillary acidic protein (GFAP) from plasma stored at - 20°C for 2, 4, 6, 15, or 35 week timepoints. This approach could simplify plasma studies and storage, particularly beneficial to enhance AD research in resource-limited settings, without significantly compromising the integrity of the biomarker measurements.

## METHODS

### Study setting and ethical considerations

The study was conducted at the John P. Hussman Institute for Human Genomics (HIHG) at the University of Miami, Miller School of Medicine. The study was performed on the blood samples obtained from ten anonymous healthy volunteers. Appropriate study ethical approval was obtained from the University of Miami ethical review board, and all the volunteers gave a written consent before the samples were collected.

### Sample collection and handling

Peripheral blood samples were collected from ten volunteers via venipuncture using a 21G needle and collected into ethylene diamine tetraacetic acid (EDTA) tubes. Plasma was isolated within 15 minutes of venipuncture via centrifugation for 10 min at 1,800 × g at room temperature (RT) and 250mL plasma aliquots were made into prelabeled barcoded polypropylene storage tubes. Two aliquots per participant were stored within 60 minutes at - 80°C and the others at -20°C for 2, 4, 6, and 15 weeks before transfer to -80°C and biomarker analysis (Figure 1). A subset of four individuals had enough plasma aliquots to extend storage to 35 weeks before transfer to -80°C.

**Figure 1:**
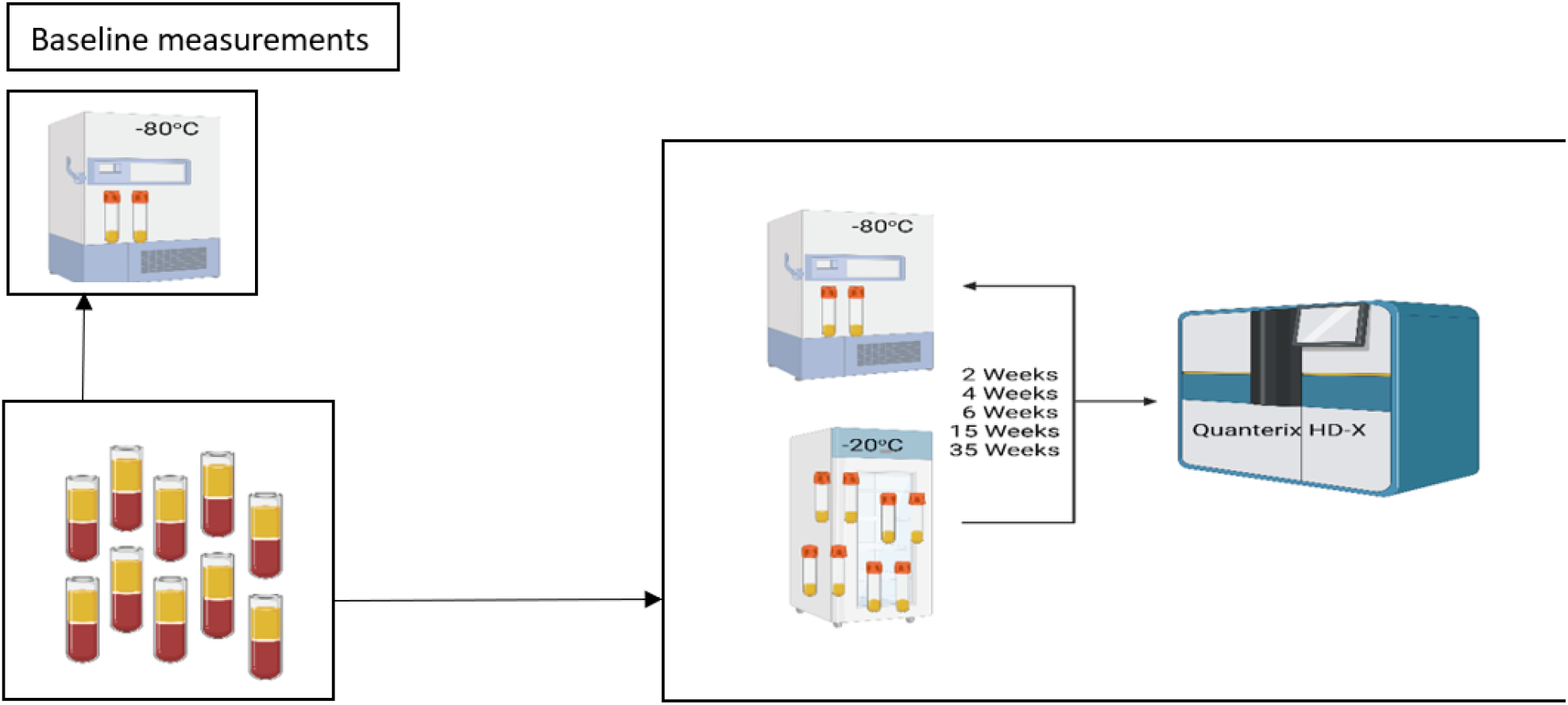
Plasma sampling, processing, and biomarkers analysis workflow.

### Sample analysis

Measurement of BBM was performed for a baseline measure from the aliquot stored at -80°C and from those stored for 2, 4, 6, 15, and 35 weeks before transfer from -20°C to -80°C. We then measured concentration of pTau181, Aβ42, Aβ40, NfL, and GFAP with Single molecule arrays (Simoa™) chemistry using the pTau181 AdvantageV2 and NEUROLOGY 4-PLEX E assays on the Quanterix HD-X [19]. Initial data analysis was performed using the Quanterix Analyzer v1.6 software to calculate standard curves and biomarker concentrations. Each aliquot was run in duplicate on the assay plate.

### Statistical analysis

The percentage change of plasma AD biomarker levels at 2, 4, 6, 15, and 35 weeks compared to the baseline were calculated for each individual. The aggregate mean value and percentage changes of each of the biomarkers at different timepoint were compared with the mean value at the baseline for each of the study participants. We considered percentage change of +/- 10 % as a cutoff point to define clinically significant changes for pTau181, Aβ42, and Aβ40 as previously suggested [17]. Variability for NfL and GFAP can be upwards of 10% within a run for the same sample as reported, suggesting that higher cutoff points are applicable for these assays (Simoa^®^ Neurology 4-Plex E).

## RESULTS

Figure 2 summarizes the percentage changes of the mean values of each biomarker assayed over time stored at -20°C before transfer to -80°C. There was a minimal mean percentage changes for pTau181 compared to the baseline from weeks 2 through 15, with mean change of <10% over the four timepoints with a noticeable decline of nearly 33% in aliquots stored for 35 weeks. The average level of Aβ42/Aβ40 ratio was stable over all study timepoints showing an overall 5% reduction compared to the baseline. However, the individual measures of Aβ42 and Aβ40 showed higher variability at the 15-week and 35-week timepoints.

**Figure 2:**
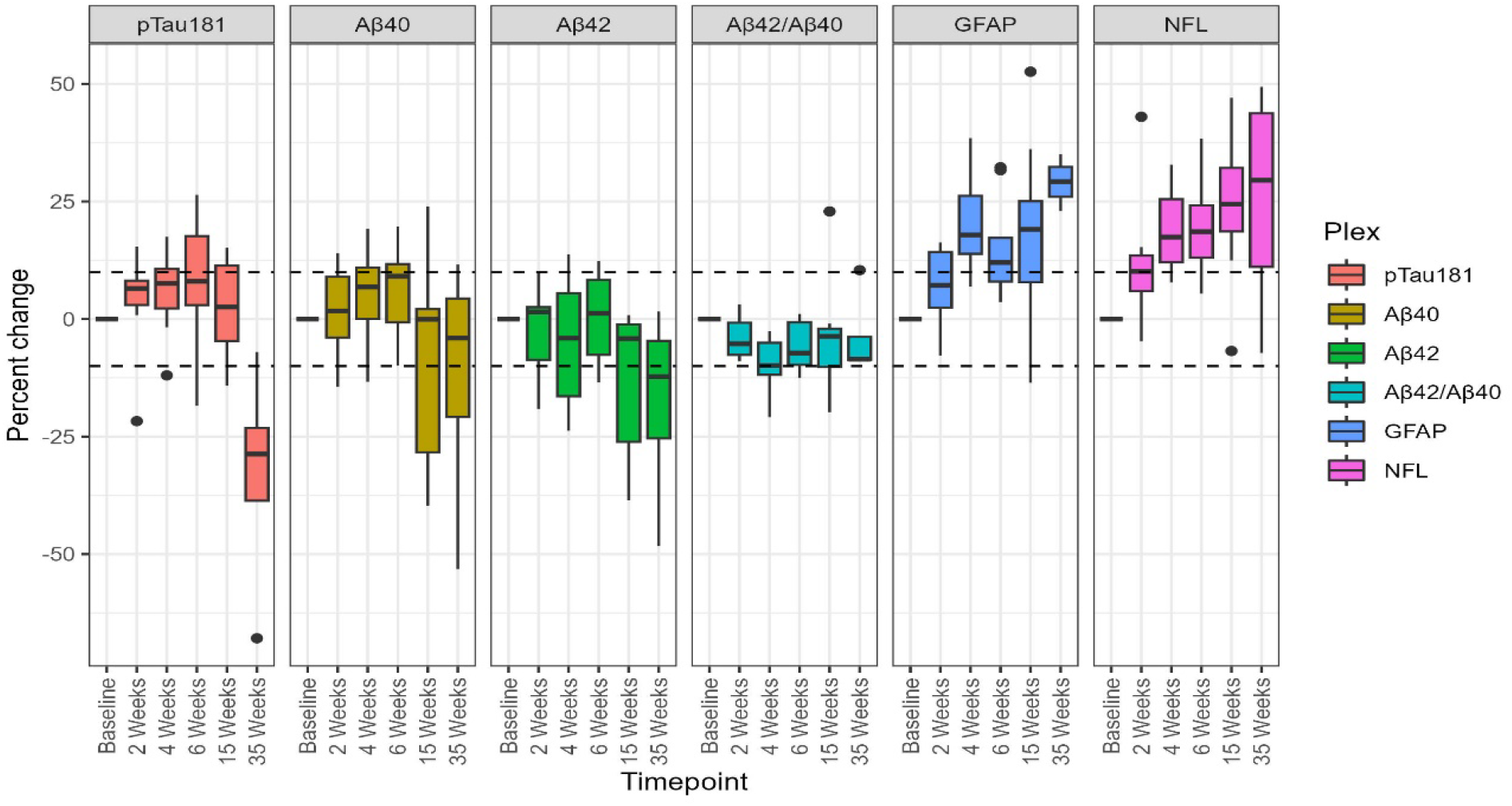
Box plots showing percentage changes of the mean of the 10 individuals of different AD blood biomarkers across five timepoints compared to the baseline. *Single solid dash line at 0: baseline measurement; *continuous dash lines: +/- 10% threshold. Black dots: outlier values for each biomarkers.

Higher variability was observed in the concentration of GFAP and NfL at all five timepoints. We observed a consistent increase in concentration of NfL at all timepoints compared to the baseline, ranging from 11% increment at week 2 to 25% increment at week 35 compared to the baseline measurement. Similar variability was observed in the level of GFAP over the five timepoints. The overall variability of GFAP ranged from 7% at week 2 to 29% at week 35 compared to the baseline value.

We have analyzed the observed variation by individual samples at various timepoints to understand the individual level variations. The concentration of pTau181 was stable in over the five time points in 8 of the study participants. In two of the samples (202301377 and 202301372) pTau181 showed greater than 10% variability at week 2, week 6, and week 35 respectively. Aβ42 and Aβ40 showed relatively consistent stability in concentration in all 10 samples at all timepoints. GFAP and NfL concentration show variations across all samples with a general increase over time (Figure 3).

**Figure 3:**
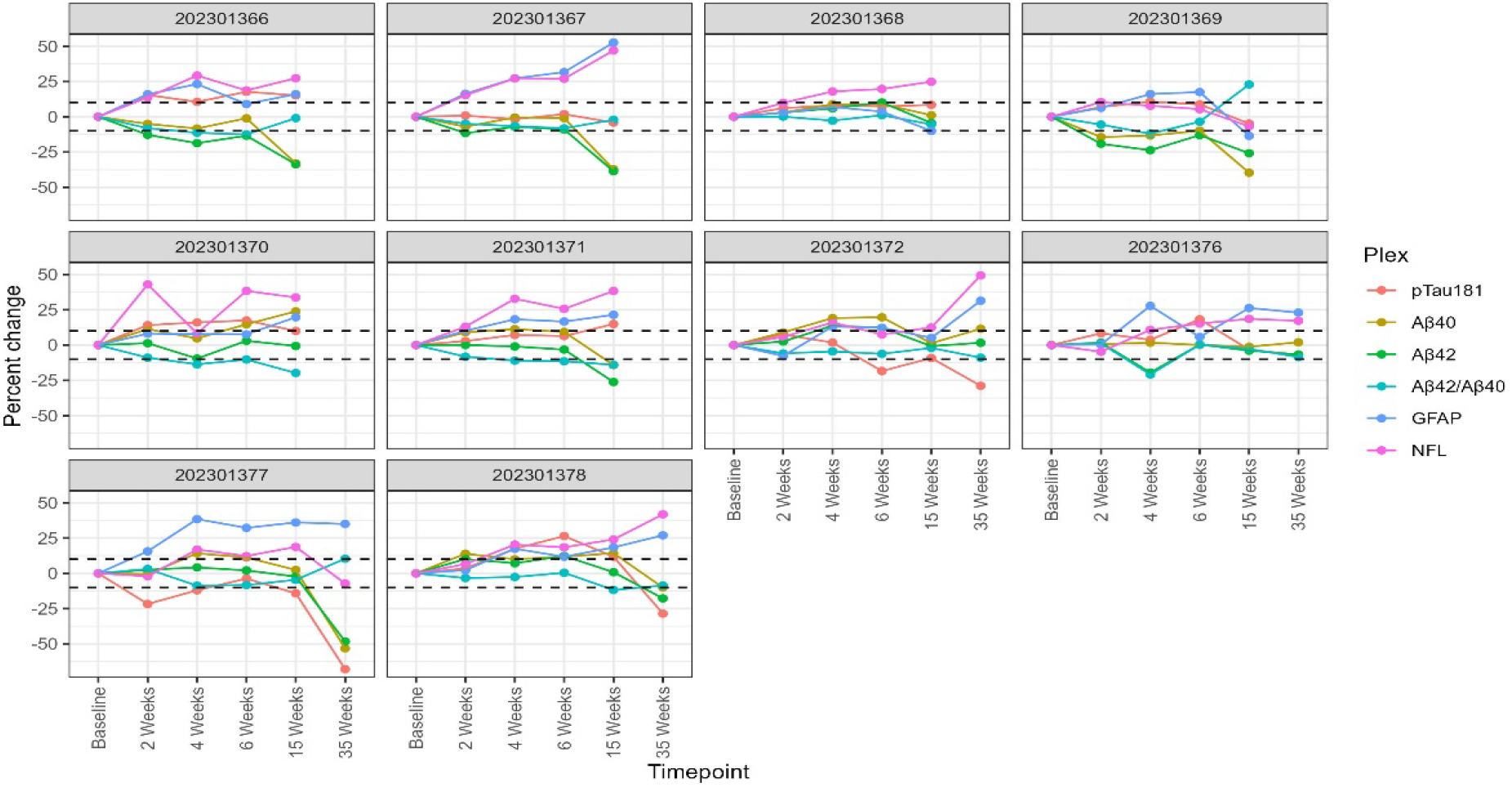
Line graph showing percentage changes of plasma biomarkers individual participants across various timepoints.

## DISCUSSION

The investigation of Alzheimer’s disease plasma biomarkers among diverse ancestry populations has been extremely limited, though increasingly studies including African Americans and Hispanic/Latino individuals from the US have emerged[20–25]. In particular, there are nearly no studies in sub-Saharan Africa, with one recent study from the Democratic Republic of Congo (DRC) a notable exception [26]. At least one barrier to inclusion of African participants in biomarker study has been the availability of research infrastructure to support the existing best practices [17]. Africa, and other low and middle income countries often lack - 80°C freezer storage due to cost of the instrumentation, reliability of the power grid, and cost of backup and solar power. Furthermore, logistics for short term storage and shipping to central repositories are hindered by lack of shipping infrastructure and availability of cold storage materials such as dry ice or liquid nitrogen. Thus, this study investigated the stability of AD BBM from plasma stored at -20°C for at timepoints beyond the current recommendation of 2 weeks. Overall, after 15 weeks of storage at -20°C, the mean levels of pTau181 and the Aβ42/Aβ40 ratio showed minimal variation compared to baseline, indicating their reliability for analysis in these settings. However, the level of pTau181 dropped significantly at week 35. Moreover, the mean concentrations of NfL and GFAP showed greater variability and increased over time points, although the overall deviations of NfL and GFAP remained within acceptable manufacturer limits, i.e. as much as 10% variability in duplicate runs.

To date, no study has investigated the stability of BBM past two weeks at -20°C freezer. Verberk et al 2022, has reported concentration of pTau181 is resistance to delayed centrifugation (24hrs) and storage at higher temperature (−20°C) for up to two weeks [17]. Despite the observed drop in the level of pTau181 at week 35, our observation supports the stability of pTau when stored in -20°C freezer up to week 15, making its usability practical in resource limited setting where the availability of a deep freezer is limited. Similarly, the Aβ42/Aβ40 ratio was stable over the measured timepoints with mild reduction from the baseline concentration. Finally, our study showed fluctuations in the concentrations of GFAP and NfL across all five measurement timepoints when compared to the baseline levels. Both the level of GFAP and NfL start increasing in the second week and maximal change from the baseline was observed at week 35.

In the present study, the overall mean concentration of pTau181 and the Aβ42/Aβ40 ratio were comparable to a recent study by Griswold et al. 2024, [24] published on MedRxiv. The study examined AD blood biomarker profiles in 2,086 individuals from diverse genetic backgrounds, aiming to identify biomarker variations among different populations. Likewise, the mean pTau181 levels were 1.98 pg/mL and 2.02 pg/mL for African Americans, and 1.84 pg/mL for Caribbean Hispanics. Additionally, they reported mean Aβ42/Aβ40 ratios of 0.0477 and 0.0575 for African Americans and 0.0432 for Caribbean Hispanics. These findings closely align with the aggregate mean concentrations observed in our study [24].

The underlying biological, chemical, or physical factors driving these deviations remain unknown and have varied across studies. Our finding is congruent with a study by Lewczuk et al. 2018, which reported the concentration of NfL increases when samples are stored even for five days at higher temperature such as room temperature or at 4 degrees [27]. However, Verbeck et al 2022, has reported they haven’t seen such an increase in level of NfL after two weeks storage in refrigerator. Furthermore, Altmann et al. 2020 reported the serum level of NfL is resistant to delayed centrifugation (24hrs) and up to three thawing and re-freezing cycles [28]. Whether these variabilities are due to biological factors such as protein aggregation, chemical factors such as assay performances, or physical features such as binding to sides of tubes remains to be investigated.

In summary, up to 15 weeks of storage of plasma at -20°C does not significantly change AD related plasma biomarker concentrations beyond assay specifications, though there are modest increases in GFAP and NfL in all timepoints. This suggests that locations lacking -80°C storage can utilize -20°C equipment and still maintain biomarker fidelity, allowing for greater representation of biomarker studies from low- and middle-income countries, increasing the diversity of studies in AD.

## Data Availability

All data produced in the present study are available upon reasonable request to the authors.

## Acknowledgement

The authors are grateful to all the ten volunteers who donated their blood to this study.

## Consent to publish

Not applicable.

## Data Availability

The raw dataset used for this study’s conclusion can be accessed upon reasonable request from the corresponding and senior author.

There are no sources in the current document.

